# Metagenome-wide association study of gut microbiome features for myositis

**DOI:** 10.1101/2021.12.15.21267821

**Authors:** Yimin Li, Jun Xu, Zijun Li, Yixue Guo, Xiaoyan Xing, Shixiong Cao, Yunzhi Zhufeng, Ziye Wang, Ruoxi Li, Dan Lu, Xu Liu, Jing He, Yuhui Li, Xiaolin Sun

## Abstract

**Objective:** The clinical relevance and pathogenic role of gut microbiome in both myositis and its associated interstitial lung disease (ILD) are still unclear. The purpose of this study was to investigate the role of gut microbiome in myositis through comprehensive metagenomic-wide association studies (MWAS).

**Methods:** We conducted MWAS of the myositis gut microbiome in a Chinese cohort by using whole-genome shotgun sequencing of high depth, including 30 myositis patients and 31 healthy controls (HC). Among the myositis patients, 11 developed rapidly progressive interstitial lung disease (RP-ILD) and 10 had chronic ILD (C-ILD). Our MWAS consisted of both overall distribution level of the bacteria analysis and pathway analysis. Receiver operating characteristic curve (ROC) analysis was performed to identify novel gut bacterial species associated with myositis or myositis-associated RP-ILD, and to evaluate their diagnostic values.

**Results:** Apparent discrepancy in β diversities of metagenome was found in the comparison of myositis and HC, RP-ILD and C-ILD in myositis. Analysis for overall distribution level of the bacteria showed *Alistipes onderdonkii, Parabacteroides distasonis* and *Escherichia coli* were upregulated, *Lachnospiraceae bacterium GAM79, Roseburia intestinalis*, and *Akkermansia muciniphila* were downregulated in patients with myositis compared to HC. *Bacteroides thetaiotaomicron, Parabacteroides distasonis* and *Escherichia coli* were upregulated, *Bacteroides A1C1* and *Bacteroides xylanisolvens* were downregulated in RP-ILD cases compared with C-ILD cases. A variety of biological pathways related to metabolism were enriched in the myositis and HC, RP-ILD and C-ILD comparison. And in the analyses for microbial contribution in metagenomic biological pathways, we have found that *E. coli* played an important role in the pathway expression in both myositis group and myositis-associated RP-ILD group. Anti-PL-12 antibody, anti-Ro-52 antibody, and anti-EJ antibody were found to have positive correlation with bacterial diversity (Shannon-wiener diversity index and Chao1, richness estimator) between myositis group and control groups. The combination of *E. coli* and *R. intestinalis* could distinguish myositis group from Healthy controls effectively. *R. intestinalis* can also be applied in the distinguishment of RP-ILD group vs. C-ILD group in myositis paitents.

**Conclusion:** Our MWAS study first revealed the link between gut microbiome and pathgenesis of myositis, which may help us understand the role of gut microbiome in the etiology of myositis and myositis-associated RP-ILD.

## Introduction

The human gut microbiome refers to the commensal microbial community that inhabits the human body and has a significant impact on host immune system and metabolism. Microbiota dysbiosis is related to the pathogenesis of a variety of autoimmune diseases including rheumatoid arthritis, inflammatory bowel diseases, systemic lupus erythematosus, multiple sclerosis and Sjogren’s syndrome [1-4]. Idiopathic inflammatory myopathies, collectively named myositis, is an autoimmune disease connected to various systemic involvement [5]. So far, no study has reported the relationship between myositis and commensal microbiota dysbiosis. In this study, we performed metagenomic-wide association study (MWAS) [6], applying metagenomic shotgun sequencing to survey associations between the gut microbiome and myositis in 30 myositis patients and 31 healthy controls (HC). Among the myositis patients, 11 developed rapidly progressive interstitial lung disease (RP-ILD) and 10 had chronic ILD (C-ILD).We revealed the microbiota dysbiosis in myositis patients, especially the patients with RP-ILD.

## Methods

### Patient participation

We examined 30 Patients diagnosed with idiopathic inflammatory myopathies (IIM) in the department of Rheumatology and Immunology, Peking University People’s Hospital in this study. There were 11 rapidly progressive interstitial lung disease (RP-ILD) cases and 10 chronic ILD (C-ILD) cases in 30 myositis patients. Cases satisfied classification criteria suggested 2017 EULAR/ACR[7]. ILD was diagnosed by the findings of high-resolution computed tomography (HRCT), according to the International Consensus Statement of Idiopathic Pulmonary Fibrosis of the American Thoracic Society [8] and the definition proposed by Suda et al.[9]. RP-ILD was defined as a progressive deterioration of ILD within three months [10]. Exclusion criteria for both sequencing groups were as follows: (i) extreme diets (e.g., strict vegetarians), (ii) treatment with antibiotics for at least three months prior to sampling. We also examined 31 healthy controls at the Peking University People’s Hospital. Healthy controls were age- and sex-matched individuals with no personal history of immune-related diseases. The characteristics of the study population were list in Supplementary Table 1.All subjects provided written informed consent before participation.

### Statistical analysis

All statistical calculations were performed using SPSS 23.0 software for Windows. Categorical variables were expressed as frequency (percentages). Numerical data were reported as the mean ± standard or mean(minimum, maximum). Continuous data were compared by student’s t-test or the Mann-Whitney *U* test. The Fisher exact test or chi-square test was used for categorical variables. P<0.05 was considered a statistically significant difference. Receiver Operator Characteristic (ROC) analysis was performed to assess the performance of the gut metagenomic biomarkers using the “pROC” package in R software (Version 3.4.1) [11].

### Sample collection and DNA extraction

Total DNA was extracted from samples using Feces:QIAamp DNA Stool Mini Kit (QIAGEN, Hilden, Germany). According to the manufacturer’s protocols. DNA completeness and purity were checked by running the samples on 1.2% agarose gels. Qubit Fluorometer was used to check the DNA concentration.

### Metagenomic library preparation and sequencing

Extracted DNA was sheared on a Covaris M220 (Covaris, Woburn, MA, USA) programmed to generate 300-bp fragments. The sequencing libraries were constructed with a NEBNext® Ultra™ DNA Library Prep Kit for Illumina® (NEB, USA). The products were purified using AgarosAgencourt AMPure XP (Beckman, USA) and quantified using the GenNext™ NGS Library Quantification Kit (Toyobo, Japan).

The libraries were sequenced using the Illumina Novaseq 6000,150-bp paired-end technology at TinyGen Bio-Tech (Shanghai) Co., Ltd.

### Bioinformatic analysis

The raw fastq files were demultiplexed based on the index. The raw, paired-end reads were trimmed and quality controlled using trimmomatic (http://www.usadellab.org/cms/?page=trimmomatic) and kneaddata software[12]. The host sequences were removed by usingbowtie2. The clean data were assembled and predicted by the software HUMAnN2, MetaPhlan2, Kraken2, and Megahit (https://github.com/voutcn/megahit) [13] and METAProdigal (http://prodigal.ornl.gov/). The taxonomies information included Domain, Kingdom, Phylum, Class, Order, Family, Genus, Species.

To understand the functions of the differentially expressed genes, the genes were assignment against the Metacyc database by using the BLASTP (BLAST Version 2.2.28+,http://blast.ncbi.nlm.nih.gov/Blast.cgi) of the e-value of 1e-5 [14].

For the Volcano Graph, Heatmap analysis, PCoA were calculated were plotted by R software (Version 3.4.1).

## Results

### Flora characteristics in the myositis gut microbiome

We selected bacteria with an average relative abundance of >1*10^−3^ to analyse, including 9 phyla, 19 classes, 29 orders, 58 families, 145genus, 398 species. Principal co-ordinates analysis (PCoA) based on Bray Curtis distance showed that the bacteria were significantly different between the myositis cases and HC from the perspective of β diversity (adonis *P*=0.001, Figure 1A). To identify flora characteristics in the myositis gut microbiome, we compared the overall distribution of the bacteria in myositis and HC (Figure 1B). At the species level, *Alistipes onderdonkii, Parabacteroides distasonis* and *Escherichia coli* were upregulated, *Lachnospiraceae bacterium GAM79, Roseburia intestinalis*, and *Akkermansia muciniphila* were downregulated in patients with myositis compared to HC. We also identified that the majority of myositis-enriched clades belonged to *L. paracasei, E. avium, etc*. Detailed results were shown in Figure 1C. We further performed gene set enrichment analysis (GSEA) based on Metacyc databases [15] to identify metagenomic biological pathways which might be involved in the pathgenesis of myositis (Figure 1D). By GSEA analysis, we found that these genes were enriched in dTDP-N-acetylthomosamine biosynthesis, fatty acid and beta-oxidation I, L-lysine biosynthesis, L-citrulline biosynthesis, *etc*. Additionally, microbial contribution in these pathways was also evaluated. The analysis showed that *Escherichia coli* accounted for the largest relative abundence of all gut microbiome, which indicated that *E*.*coli* may play an important role in the pathway of dTDP-N-acetylthomosamine biosynthesis (Figure 1E), L-tryptophan biosynthesis (Supplementary Figure 1A), NAD biosynthesis I (from aspartate) (Supplementary Figure 1B), L-arginine biosynthesis IV (archaebacteria) (Supplementary Figure 1C), *etc*.

**Figure 1.**
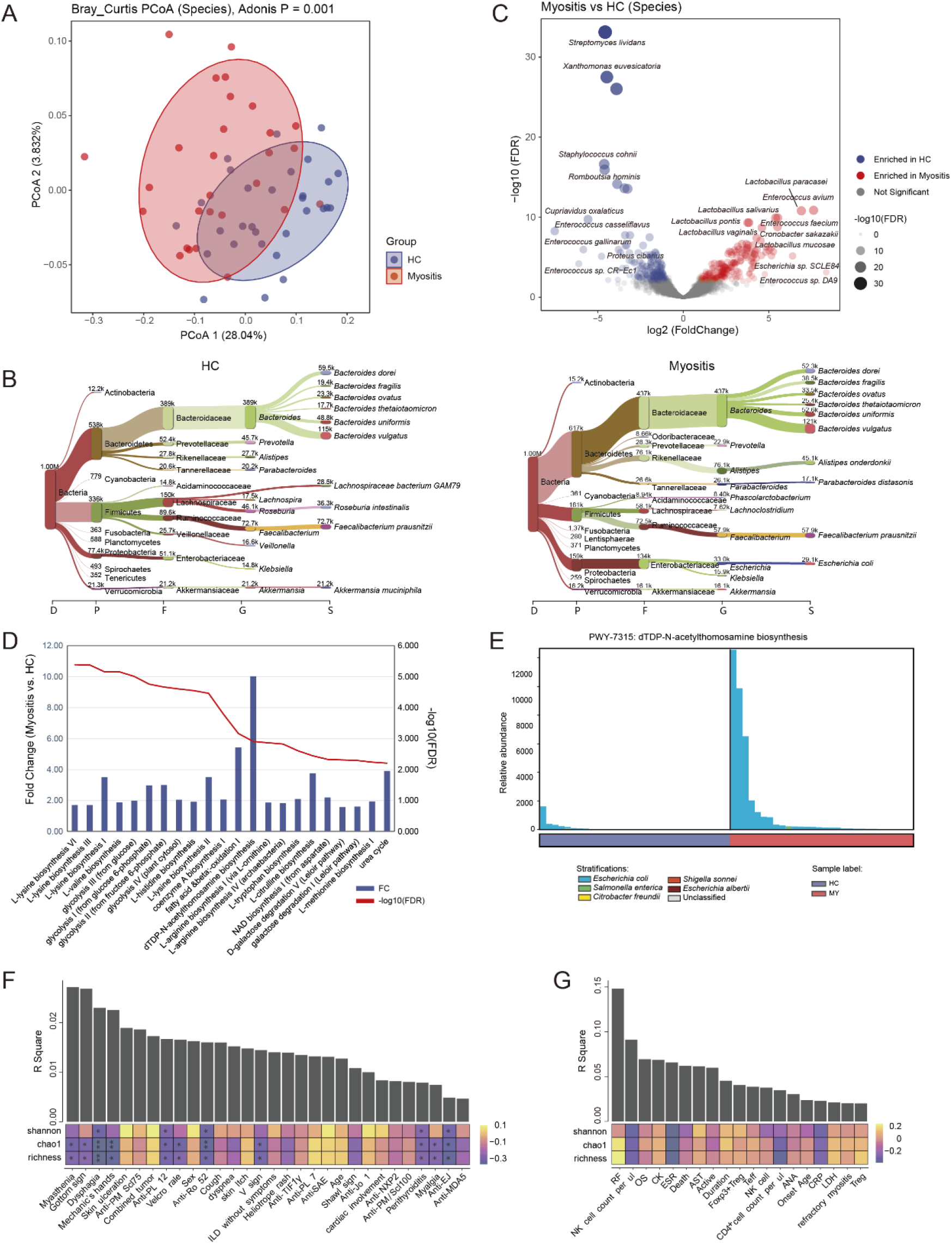
MWAS results of myositis-HC pathway association tests. (A) The principal co-ordinates analysis (PCoA) based on Bray Curtis distance. Distance matrix was evalued with the bacterial composition at the species level. The Adonis test was performed to compare the significance of beta diversity. (B) Analysis for overall distribution level of the bacteria in myositis and HC, the top 10 are listed. Visualization was performed with the online server of Pavian metagenomics data explorer (https://fbreitwieser.shinyapps.io/pavian/). (C) A volcano plot of the MWAS results of the clades. The x-axis indicates log2 Fold Change values. The y-axis indicates observed −log10 FDR values. Clades with Fold Change values more than 2 and FDR values less than 0.01 are regarded as enriched. (D) A gene set enrichment analysis based on Metacyc databases to identify metagenomic biological pathways. Blue bars showed the comparative fold change value (Myositis vs. HC), and red line presented the -logFDR value. (E) Bacterial contribution in specific biological pathway (FAO-PWY: *dTDP-N-acetylthomosamine biosynthesis*). The correlation analysis between clinic parameters (Complications, F; immune indexes, G) and bacterial beta/alpha diversity. The bar represented the R square value of the correlation between clinic indexes and bacterial beta diversity, and pheatmap showed the Spearman’s correlation between clinic indexes and alpha diversity (Shannon-wiener diversity index and Chao1, richness estimator). *FDR<0.05, **FDR <0.01, ***FDR<0.001. HC: healthy controls; MY: myositis; OS: overall survival; ANA: anti-nuclear antibody; TIF-1γ: translation initiation factor-1γ; MDA5: melanoma differentiation-associated 5; NXP2: nuclear matrix protein 2; SAE: small ubiquitin-like modifier enzyme; PM/Scl: polymyositis/scleroderma; RF: rheumatoid factor; AST: aspartate aminotransferase; LDH: lactate dehydrogenase; CK: creatine kinase; ESR: erythrocyte sedimentation rate; CRP: C-reactive protein; NK: natural killer cell; Treg: Regulatory T cells; Teff: effector T cells.

The presence of anti-PL-12 antibody, anti-Ro-52 antibody, or anti-EJ antibody was significantly correlated with decreased bacterial diversity (Shannon-wiener diversity index and Chao1, richness estimator), while anti-PL-7 antibody, anti-SAE antibody and age showed positive correlation with bacterial diversity (Figure 1F-G). We compared the gut microbiota of ASS patients and dermatomyositis (DM) patients in this myositis cohort to investigate the difference between them. β diversity analysis showed no significant gut microbiome difference between the ASS patients and the DM patients (Supplementary Figure 2). The bigger R-square shows the better correlation, so myasthenia and RF had more significant correlation than other clinical characters among them(Figure 1F-G).

These data revealed significant differences in the gut microbiome and related metagenomic biological pathways between myositis patients and HC, which might be involved in the pathogenesis of myositis.

### Identification of gut microbiome associated with RP-ILD in myositis

Interstitial lung disease (ILD) is an important complication of myositis, which can be divided into rapidly progressive ILD (RP-ILD) and chronic ILD (C-ILD). RP-ILD is the most fatal complication for myositis [16]. In order to explore the association between gut microbiota dysbiosis and ILD development in myositis patients, we divided the myositis cases into three groups: RP-ILD, C-ILD and Non-ILD. β diversity analysis showed a difference in mibrobiota diverstiy between the RP-ILD group and the C-ILD group (adonis *P*=0.029, Figure 2A), but no significant difference was found between patients with ILD and without ILD (Supplementary Figure 3).

**Figure 2.**
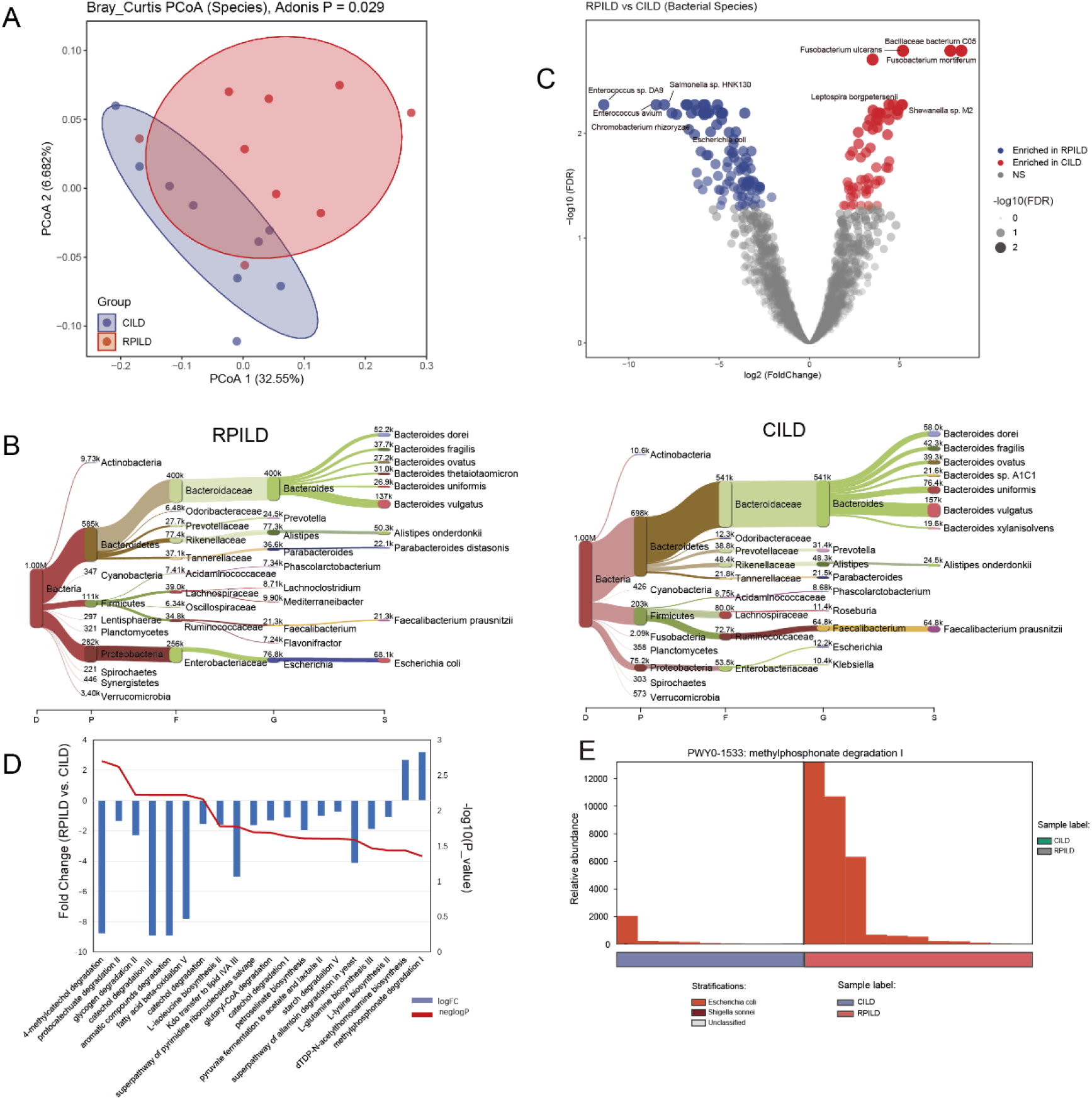
MWAS results of myositis-associated RP-ILD andC-ILD pathway association tests. (A) The principal co-ordinates analysis (PCoA) based on Bray Curtis distance. Distance matrix was evalued with the bacterial composition at the species level. The Adonis test was performed to compare the significance of beta diversity. (B) Analysis for overall distribution level of the bacteria in RP-ILD and C-ILD, the top 10 are listed. Visualization was performed with the online server of Pavian metagenomics data explorer (https://fbreitwieser.shinyapps.io/pavian/). (C) A volcano plot of the MWAS results of the clades. The x-axis indicates log2 Fold Change values. The y-axis indicates observed ™log10 FDR values. Clades with Fold Change values more than 2 and FDR values less than 0.01 are regarded as enriched. (D) A gene set enrichment analysis based on Metacyc databases to identify metagenomic biological pathways. Blue bars showed the comparative fold change value (RP-ILD vs. C-ILD), and red line presented the -logFDR value. (E) Bacterial contribution in specific biological pathway (FAO-PWY: *methylphosphonate degradation I*). RP-ILD: rapidly progressive interstitial lung disease; C-ILD: chronic interstitial lung disease.

To compare microbial characteristics between the RP-ILD group and C-ILD group, we also analyzed the overall distribution level of the bacteria. The results were shown in Figure 2B. At the level of Species, *Bacteroides thetaiotaomicron, P. distasonis* and *E. coli* were upregulated, *Bacteroides A1C1* and *Bacteroides xylanisolvens* were downregulated in RP-ILD cases compared with C-ILD cases. Next, we further identified that the majority of RP-ILD-enriched clades belonged to *E. aviu, E. sp*.*DA9, etc*. Detailed results are shown in Figure 2C. GSEA was performed to identify metagenomic biological pathways associated with RPILD (Figure 2D). By GSEA analysis, we found that enriched pathways were 4-methylcatechol degradation, fcatechol degradation, aromatic compounds degradation, fatty acid beta-oxidation V, methylphosphonate degradation I and *ect*. Additionally, microbial contribution in these pathways was also evaluated. By GSEA analysis, these genes were found to be enriched in methylphosphonate degradation I, dTDP-N-acetylthomosamine biosynthesis, starch degradation V, and *ect*, with the result as *E*.*coli* made up the largest relative abundence proportion in all gut microbiome, which implied *E*.*coli* may play an considerable part in the pathway of methylphosphonate degradation I (Figure 2E), dTDP-N-acetylthomosamine biosynthesis (Supplementary Figure 4A), starch degradation V (Supplementary Figure 4B).

These data revealed significant differences in the gut microbiome and metagenomic biological pathways between myositis patients with RP-ILD and C-ILD, implicating differential gut microbiome might be involved in development of RP-ILD in myositis.

### The diagnostic potential of microbial contents for myositis and myositis-associated RP-ILD

According to the results of the gut microbiome screening, we performed ROC analysis to evaluate their diagnostic values in both grouping-situation: myositis group vs. control groups, RP-ILD group vs. C-ILD group in myositis patients. We selected two bacteria with high area under curve (AUC) value in many common different gut microbiome, including *E. coli* and *R. intestinalis*, for analysis. In the above analyses for microbial contribution in metagenomic biological pathways, we have found that *E. coli* played an essential role in the pathway expression in both the myositis group and RP-ILD group. Next, we found that the data of *E. coli, R. intestinalis*, and the combination of *E. coli* and *R. intestinalis* all had good diagnostic values. In myositis group vs. control groups, the AUC of *E. coli, R. intestinalis*, and the combination of *E. coli* and *R. intestinalis* were 0.705 (95%CI, 0.569-0.842), 0.860 (95%CI, 0.765-0.956) and 0.867 (95%CI, 0.773-0.960), respectively (Figure 3A). The combination of *E. coli* and *R. intestinalis* had the best diagnostic values to distinguish myositis patients from healthy controls. In RP-ILD group vs. C-ILD group, the AUC of *E. coli, R. intestinalis*, and the combination of *E. coli* and *R. intestinalis* were 0.737 (95%CI, 0.508-0.967), 0.879 (95%CI, 0.725-1.000) and 0.677 (95%CI, 0.428-0.926), respectively (Figure 3B). *R. intestinalis* alone had the best diagnostic values to distinguish myositis patients with RP-ILD from those with C-ILD. This result suggested that differences in abundance of *R*.*intestinalis* could characterize RP-ILD development in myositis patients.

**Figure 3.**
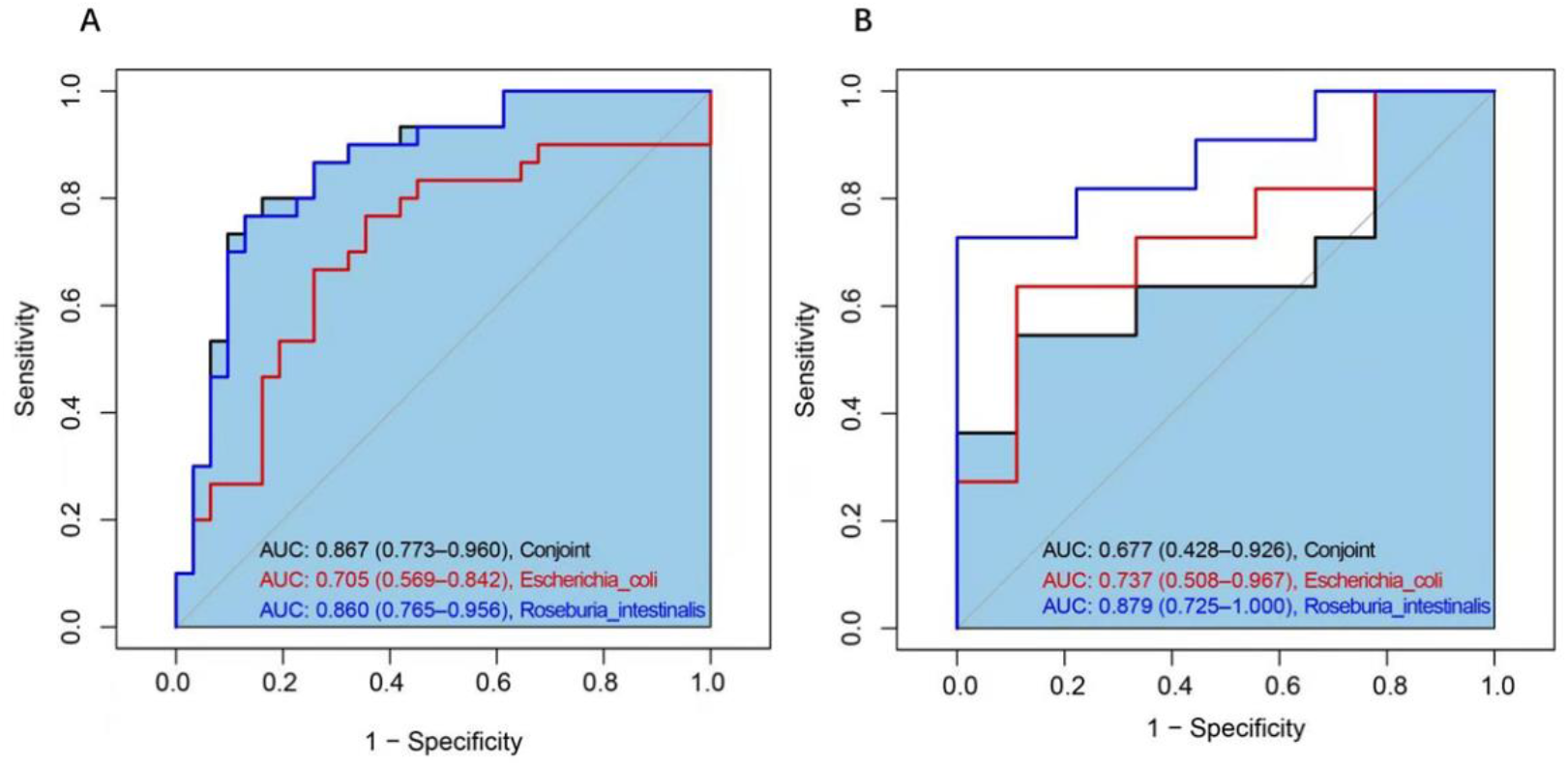
The ROC curves used for discriminating myositis from HC, and RP-ILD from C-ILD. (A) Myositis group vs. Control groups (B) RP-ILD group vs. C-ILD group.ROC: receiver operating characteristic curves.

## Disscussion

Myositis represents a heterogeneous group of autoimmune diseases featured by muscle inflammation. By performing a comprehensive MWAS analysis of the gut microbiota in the myositis case-control cohort, our study proposed the link between gut microbiome and myositis or myositis-associated ILD, and showed the signaling pathways affected by gut microbiota associated with this disease.In the comparison of myositis and HC, *A. onderdonkii, P. distasonis* and *E. coli* were upregulated in myositis, *L. bacterium GAM79, R. intestinalis*, and *A. muciniphila* were downregulated in HC. ROC analysis also demonstrated that a model of the combination of *E. coli* and *R*.*intestinalis* had good diagnostic values in myositis group vs. HC groups. *R*.*intestinalis* alone could distinguish myositis patients with RP-ILD from those with C-ILD.

Previous studies suggested that the *A. onderdonkii* seemed as a probiotic, whose reduction could contribute to the advanced fibrosis in the nonalcoholic fatty liver disease (NAFLD) patients by decreasing the production of short-chain fatty acids [17, 18]. However, in our study, it was found to be a prevalent species in the myositis group, which suggested *A. onderdonkii* might be involved in the disease development of myositis.

A recent report proved that *P. distasonis* could improve the disorder of host glucose and lipid metabolism [19], while one study reported that genes essential to glycolysis were suppressed in myositis [20]. In this study, our results showed *P. distasonis* was upregulated in myositis group, accompanied by the high expression of metagenomic biological pathways <u>o</u>f glucose and lipid metabolism, such as glycolysis I (from glucose 6-phosphate), fatty acid and beta oxidation I, etc. A previous study had reported that *P. distasonis* can alleviate obesity and metabolic dysfunctions in *ob/ob* and high-fat diet (HFD)-fed mice [19]. According to our results, we speculated *P. distasonis* might play a complicated role, having different function under different backgrounddisease conditions, and might be pathogenic in myositis.

Several reports demonstrated that Toll-like receptor (TLR)-4 and TLR3, two potent inducers of autophagy, were highly expressed in myositis compared to controls [21, 22]. *E. coli*, through its bacterial amyloid curli, can trigger the innate immunity via TLRs (including TLRs 1/2, 4, and 9) and inflammasome [23, 24], thereby constituting a potential immune trigger for individuals genetically predisposed to myositis. Moreover, curli amyloid is naturally complexed to bacterial extracellular DNA (eDNA) and make *E. coli* activate dendritic cells [25] and macrophages [26] to produce a lot of proinflammatory factors and can trigger the production of type I IFNs [27]. It is possible that enriched *E. coli* might function as a trigger of the pathogenic immune response for myositis.

In our results, both *A*.*muciniphila* and *R*.*intestinalisis* were upregulated in HC compared to myositis, and both of them were reported to be promising probiotics [28, 29]. Mucin is implicated in mucosal immune defense by protecting the surface epithelium of the airways and intestine. Increased secretion of mucin may interfere with the normal repair process of the epithelium or may be toxic and stimulate a fibroproliferative response to cause both myositis and ILD [30, 31]. Since *A. muciniphila* could degrade mucin to allievate inflammatory bowel disease (IBD) [32, 33], it might play a positive role in myositis improvement. One metabolite of *A. muciniphila*, propionic acid, might be involved in the its immunoregulatory function. *R. intestinalis* ameliorated IBD by reducing inflammatory macrophages and activated CD4^+^ cells in the colon [29] and producing short-chain fatty acids, especially butyrate, affecting colonic motility, immunity maintenance and anti-inflammatory properties [34]. Since uncontrolled inflammation is featured in the pathogenesis of myositis [30, 35], the decrease of anti-inflammatory *R*.*intestinalis* might contribute to myositis development. The relationship between myositis and *R*.*intestinalis* or *A*.*muciniphila* has not been revealed. The possible roles of *R*.*intestinalis* and *A*.*muciniphila* as probiotics [23, 24] in myositis development are worthy of further investigation. Moreover, our results showed *L*.*bacterium GAM79* was upregulated in the HC. However, no study on the function of *L*.*bacterium GAM79* has been reported.

Overall, there were no differences observed between ILD and Non-ILD groups in β diversity analysis, the result might be limited due to the small sample size after stratification in this study. But in the comparison between RP-ILD and C-ILD groups, we found *B. thetaiotaomicron, P. distasonis* and *E. coli* were upregulated, *B. A1C1* and *B. xylanisolvens* were downregulated in RP-ILD group.

Recently, one study confirmed that the levels of oxidized fatty acids in patients with myositis were significantly higher than those in HC, especially in myositis patients with ILD [36]. In our results, *P. distasonis* was also accompanied by the decreased expression of pathways about glucose and lipid metabolism in myositis-associated RP-ILD group, such as glycogen degradation III, fatty acid and beta oxidation V. This result also implicated that *P. distasonis* might affect the disease process of myositis-associated RP-ILD by down-regulating the expression of glucose and lipid metabolism pathways.*B. thetaiotaomicron, B. xylanisolvens* and *B. A1C1* are specices of *Bacteroides*, which maintain a complex and generally beneficial relationship with the host when retained in the gut [37]. *B. thetaiotaomicron* can stimulate production of an antibiotic Paneth cell protein (Ang4) that can kill certain pathogenic organisms [38], and induce Paneth cells to produce a bactericidal lectin, RegIIIγ, which exerts its antimicrobial effect by binding to the peptidoglycan of gram-positive organisms [39]. *B. thetaiotaomicron* also participates in a variety of glucose and lipid metabolism pathways [40]. According to previous studies, *B. thetaiotaomicron* seemed to be probiotic, but our results showed that *B. thetaiotaomicron* was more prevalent in the RP-ILD group associated with higher mortality, which indicated *B. thetaiotaomicron* might increase as a negative feedback factor or play a pathogenic role in RP-ILD, whose exact roles needs further research in the future.. *B. xylanisolvens* can alleviate nonalcoholic hepatic steatosis and atherosclerosis as a probiotic by increasing gut folate level and promoting folate-dependent one-carbon metabolism [41]. Moreover, it was identified as a prevalent specie in type 1 diabetes patients [42]. These results showed that *B. xylanisolvens* might play a complicated role, having different metabolic characteristics and regulating different signaling pathways under different background and growth conditions. There was no report about *B. A1C1*, and our results showed both *B. xylanisolvens* and *B. A1C1* were downregulated in RP-ILD group, which implied both of them might be probiotic in myositis-associated RP-ILD.

This is the first study to investigate the role of gut microbiome in myositis or myositis-associated ILD through MWAS. Although the biological mechanisms underlying the effects of gut microbiome, such as *P. distasonis, B. thetaiotaomicron*, are still unclear, our data indicate profound influences of these microbiome in the pathogenesis of myosits or myositis-associated ILD. In conclusion, our study provided a number of myositis-associated bacterial species representing potential microbial targets for future functional research to further clarify the roles of the microbiome in the etiology of myositis.

## Data Availability

All data produced in the present study are available upon reasonable request to the authors

## Patient and public involvement

Patients and/or the public were not involved in the design, or conduct, or reporting, or dissemination plans of this research.

## Patient consent for publication

Not required.

## Ethics approval

This study was approved by the ethics committee of Peking University People’s Hospital (Document ID:2020PHB114-01).

---

**Supplementary Figure 1.**
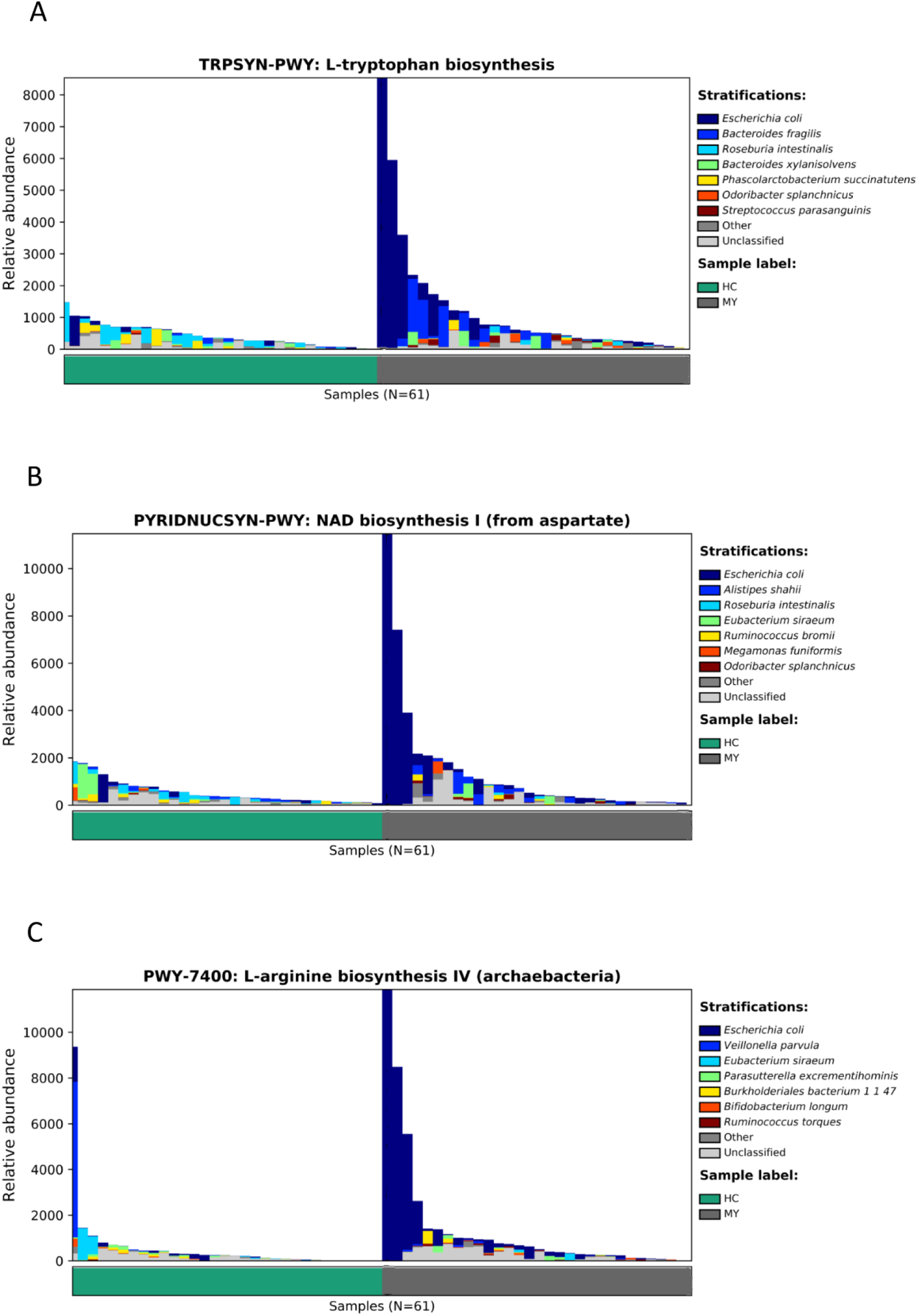
Bacterial contribution in specific biological pathway :FAO-PWY: *L-tryptophan biosynthesis* (A), *NAD biosynthesis I (from aspartate)* (B), *L-arginine biosynthesis IV (archaebacteria)* (C). HC: healthy controls; MY: myositis.

**Supplementary Figure 2.**
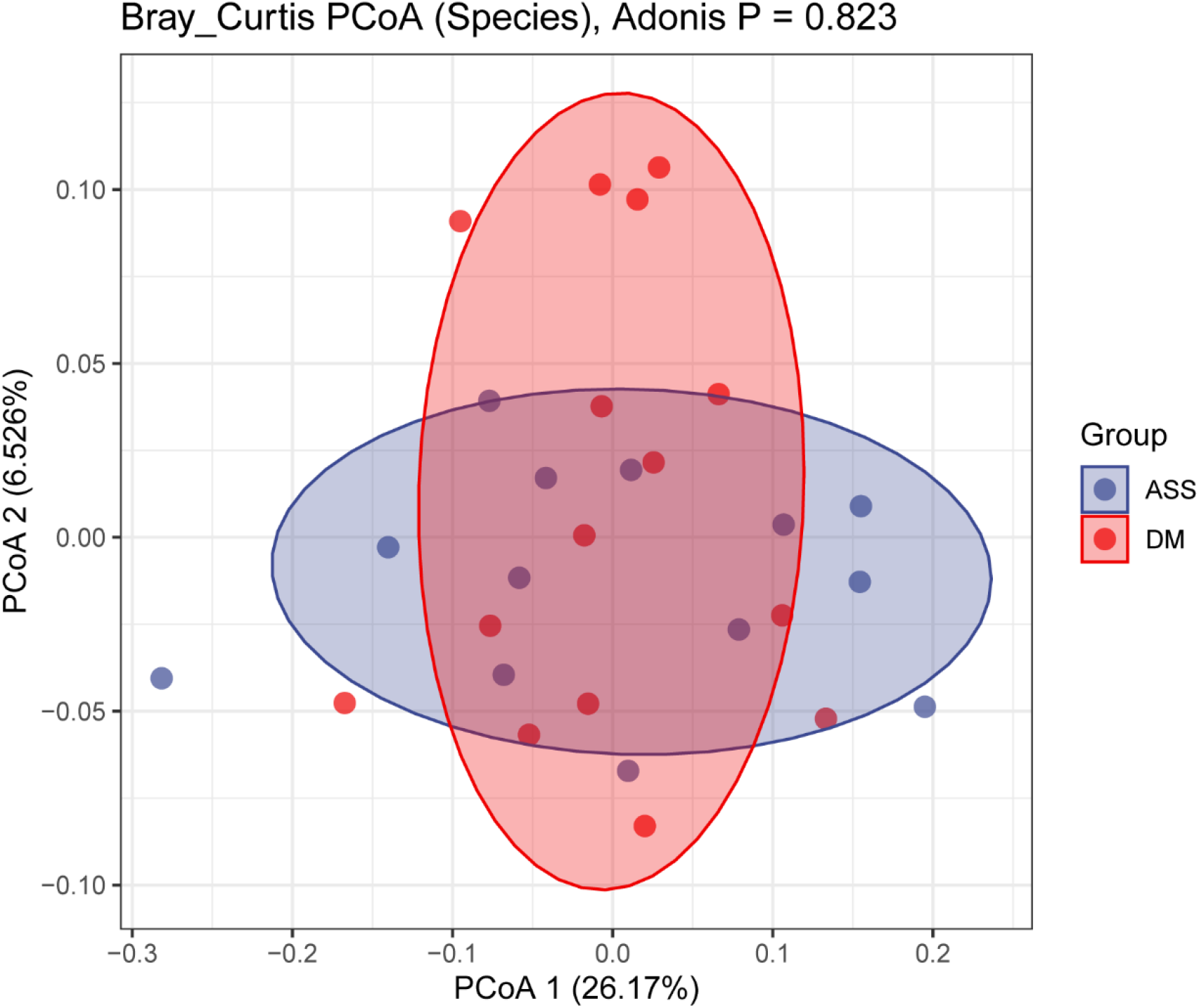
The principal co-ordinates analysis (PCoA) between ASS and DM based on Bray Curtis distance. Distance matrix was evalued with the bacterial composition at the species level. The Adonis test was performed to compare the significance of beta diversity.ASS: anti-synthetase syndrorme;DM: dermatomyositis.

**Supplementary Figure 3.**
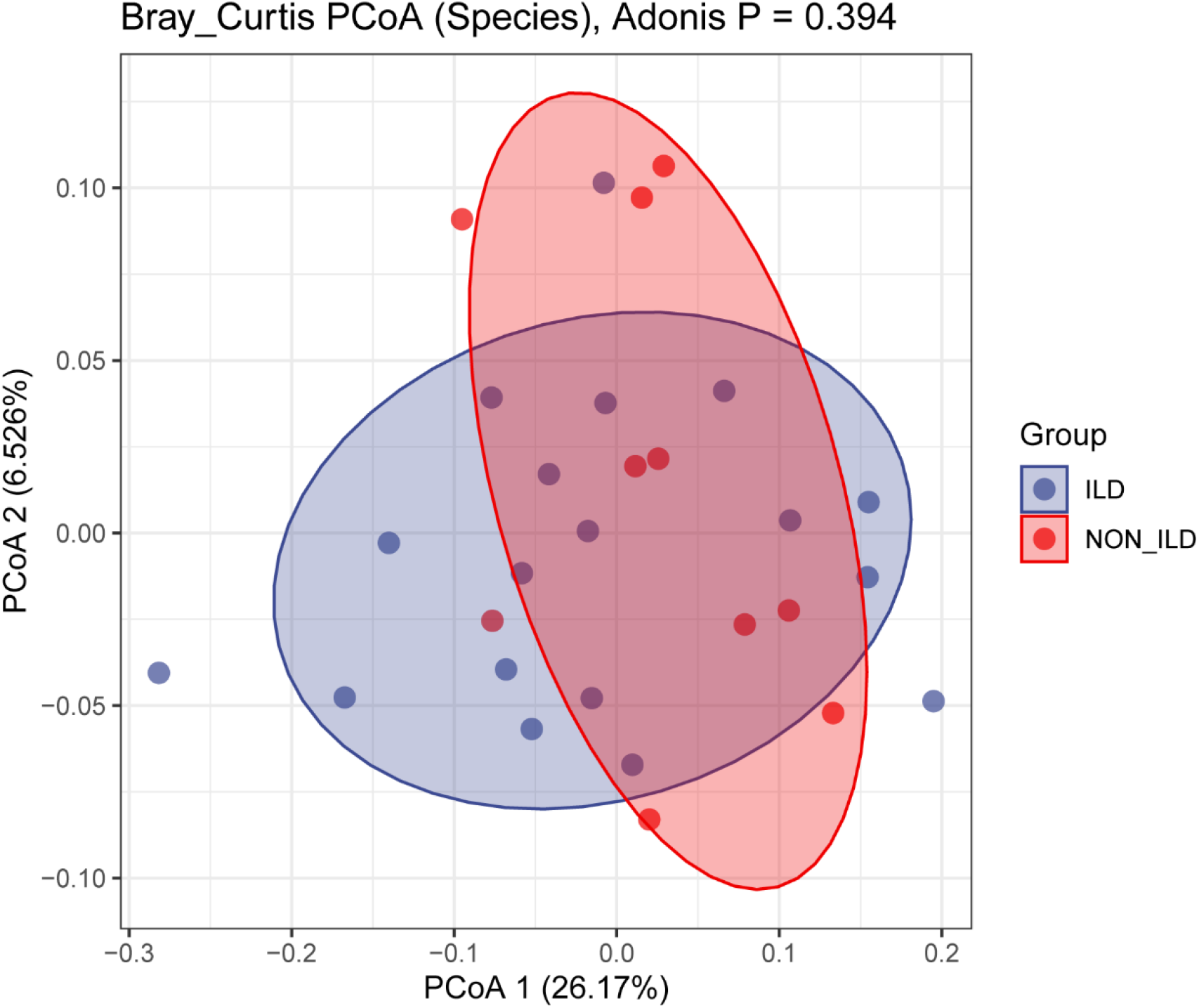
The principal co-ordinates analysis (PCoA) between ILD and Non-ILD based on Bray Curtis distance. Distance matrix was evalued with the bacterial composition at the species level. The Adonis test was performed to compare the significance of beta diversity.ILD: interstitial lung disease.

**Supplementary Figure 4.**
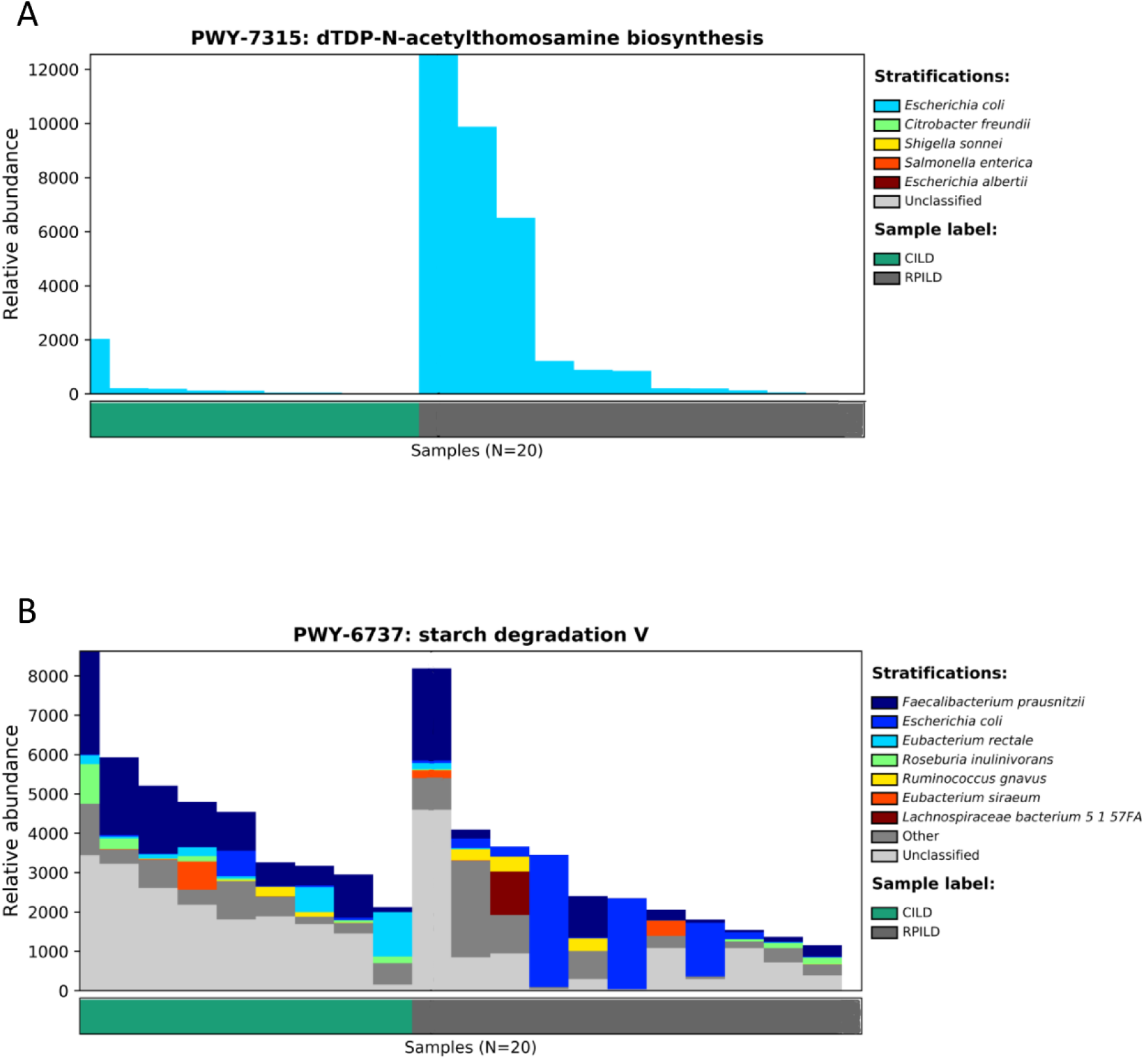
Bacterial contribution in specific biological pathway :FAO-PWY: *dTDP-N-acetylthomosamine biosynthesis* (A), *starch degradation V* (B). RP-ILD: rapidly progressive interstitial lung disease; C-ILD: chronic interstitial lung disease.

**Supplementary Table 1.** Comparison of clinical and laboratory characteristics between patients with myositis and HC.

|  | Myositis (n=30) | HC (n=31) | *P* value |
| --- | --- | --- | --- |
| Age, years | 50.7±15. | 55.6±5.0 | 0.371 |
| Age at onset, years | 49.6±16.2 | / | / |
| Duration of rash, months | 17.7(0.5,108) | / | 0.000 |
| OS, years | 1.74(0,9) | / | / |
| Death | 3 (10) | 0 (0) | 0.073 |
| Female | 21 (70) | 24 (77.4) | 0.581 |
| Gotttron | 24 (80) | 0 (0) | 0.000 |
| Mechanic hand | 19 (63.3) | 0 (0) | 0.000 |
| heliotrope rash | 11 (36.7) | 0 (0) | 0.000 |
| V sign | 14 (46.7) | 0 (0) | 0.000 |
| Shawl sign | 12 (40) | 0 (0) | 0.000 |
| Skin ulceration | 2 (6.7) | 0 (0) | 0.149 |
| Skin Itch | 4 (13.3) | 0 (0) | 0.036 |
| Perithyroiditis | 8 (26.7) | 0 (0) | 0.002 |
| Myalgia | 13 (43.3) | 0 (0) | 0.000 |
| Myasthenia | 17 (56.7) | 0 (0) | 0.000 |
| Cough | 13 (43.3) | 0 (0) | 0.000 |
| Dyspnea | 11 (36.7) | 0 (0) | 0.000 |
| Velcro rale | 11 (36.7) | 0 (0) | 0.000 |
| ILD without symptoms | 13 (43.3) | 0 (0) | 0.000 |
| Combined tumor | 1 (3.3) | 0 (0) | 0.313 |
| MSAs/MAAs |  |  |  |
| ANA | 26 (86.7) | 0 (0) | 0.000 |
| Anti-TIF-1γ | 1 (3.3) | 0 (0) | 0.313 |
| Anti-MDA5 | 8 (26.7) | 0 (0) | 0.002 |
| Anti-NXP2 | 2 (6.7) | 0 (0) | 0.149 |
| Anti-SAE | 3 (10) | 0 (0) | 0.313 |
| Anti-PM/Scl100 | 1 (3.3) | 0 (0) | 0.313 |
| Anti-PM/Scl75 | 2 (6.7) | 0 (0) | 0.149 |
| Anti-Jo-1 | 5 (16.7) | 0 (0) | 0.017 |
| Anti-PL-7 | 1 (3.3) | 0 (0) | 0.313 |
| Anti-PL-12 | 5 (16.7) | 0 (0) | 0.017 |
| Anti-EJ | 3 (10) | 0 (0) | 0.073 |
| Anti-Ro52 | 14 (46.7) | 0 (0) | 0.000 |
| RF (U/L) | 32.4(0,473) | / | / |
| Cardiac involvement | 5 (16.7) | 0 (0) | 0.017 |
| Dysphagia | 4 (13.3) | 0 (0) | 0.036 |
| Disease active status | 19 (63.3) | / | / |
| Refractory myositis | 6 (20) | / | / |
| AST (U/L) | 70.4(15,316) | / | / |
| LDH (U/L) | 429.9(168,1520) | / | / |
| CK (U/L) | 828.2(20,7993) | / | / |
| ESR (mm/h) | 24.1±16.4 | / | / |
| CRP (mg/L) | 10.1(0,64) | / | / |
| Lymphocyte subsets in peripheral blood |  |  |  |
| NK cell,% | 10.5±7.7 | / | / |
| NK cell count/ul | 164.7±151.1 | / | / |
| CD4+ cell count/ul | 654.6±414.7 | / | / |
| Treg, % | 9.5±4.6 | / | / |
| Foxp3+Treg, % | 8.64±3.9 | / | / |
| Teff, % | 90.9±3.9 | / | / |

HC: healthy controls; OS: overall survival; MSAs: myositis-specific autoantibodies; MAAs: myositis-associated autoantibodies; ANA: anti-nuclear antibody; TIF-1γ : translation initiation factor-1γ ; MDA5: melanoma differentiation-associated 5; NXP2: nuclear matrix protein 2; SAE: small ubiquitin-like modifier enzyme; PM/Scl: polymyositis/scleroderma; RF: rheumatoid factor; AST: aspartate aminotransferase; LDH: lactate dehydrogenase; CK: creatine kinase; ESR: erythrocyte sedimentation rate; CRP: C-reactive protein; NK: natural killer cell; Treg: Regulatory T cells; Teff: effector T cells.

**Supplementary Table 2.** Comparison of clinical and laboratory characteristics between patients with RPILD and CILD.

|  | RPILD(n=11) | CILD(n=9) | *P* value |
| --- | --- | --- | --- |
| Age, years | 54.6±13.0 | 43±13.5 | 0.066 |
| Age at onset, years | 54.4±13.2 | 41.6±12.5 | 0.049 |
| Duration of rash, months | 1.7±1.1 | 28.6±23.7 | 0.000 |
| OS, years | 0.5(0,3) | 4.11(0,9) | 0.000 |
| Death | 3(27.3) | 0(0) | 0.098 |
| Female | 9(81.8) | 6(66.7) | 0.463 |
| Gotttron | 7(63.6) | 7(77.8) | 0.518 |
| Mechanic hand | 7(63.6) | 7(77.8) | 0.518 |
| heliotrope rash | 5(45.5) | 3(33.3) | 0.605 |
| V/sign | 4(36.4) | 3(33.3) | 0.895 |
| shawl sign | 4(36.4) | 2(22.2) | 0.518 |
| Skin ulceration | 2(18.2) | 0(0) | 0.196 |
| Skin Itch | 1(9.1) | 2(22.2) | 0.440 |
| Perithyroiditis | 3(27.3) | 1(11.1) | 0.395 |
| Myalgia | 5(45.5) | 3(33.3) | 0.605 |
| Myasthenia | 5(45.5) | 3(33.3) | 0.605 |
| Cough | 7(63.6) | 4(44.4) | 0.418 |
| dyspnea | 9(81.8) | 1(11.1) | 0.001 |
| Velcro rale | 10(90.9) | 1(11.1) | 0.000 |
| ILD without symptoms | 8(72.7) | 3(33.3) | 0.086 |
| Combined tumor | 0(0) | 0(0) | / |
| MSAs/MAAs |  |  |  |
| ANA | 9(81.8) | 9(100) | 0.196 |
| Anti-TIF-1γ | 0(0) | 0(0) | / |
| Anti-MDA5 | 7(63.6) | 0(0) | 0.002 |
| Anti-NXP2 | 0(0) | 0(0) | / |
| Anti-SAE1 | 0(0) | 0(0) | / |
| Anti-PM/Scl100 | 0(0) | 1(11.1) | 0.281 |
| Anti-PM/Scl75 | 1(9.1) | 1(11.1) | 0.834 |
| Anti-Jo-1 | 2(18.2) | 3(33.3) | 0.665 |
| Anti-PL/7 | 0(0) | 1(11.1) | 0.281 |
| Anti-PL/12 | 2(18.2) | 1(11.1) | 0.769 |
| Anti-EJ | 1(9.1) | 2(22.2) | 0.406 |
| Anti-Ro52 | 5(45.5) | 6(66.7) | 0.483 |
| RF (U/L) | 2.84(0,27) | 54.84(0,473) | 0.826 |
| Cardiac involvement | 1(9.1) | 1(11.1) | 0.888 |
| Dysphagia | 1(9.1) | 0(0) | 0.380 |
| Disease active status | 11(100) | 4(44.4) | 0.002 |
| Refractory myositis | 4(36.4) | 1(11.1) | 0.215 |
| AST (U/L) | 85.0(15,316) | 36.9(19,101) | 0.376 |
| LDH (U/L) | 486.4(202,1520) | 292.7(168,628) | 0.103 |
| CK (U/L) | 367.7(21,3022) | 871.9(20,3163) | 0.103 |
| ESR (mm/h) | 25.7±18.0 | 25.2±17.2 | 0.913 |
| CRP (mg/L) | 8.38(0,44) | 15.77(0,64) | 0.440 |
| Lymphocyte subsets in peripheral blood |  |  |  |
| NK cell,% | 8.8±8.4 | 11.3±8.6 | 0.524 |
| NK cell count/ul | 151.8 (0,620) | 148.4(57,386) | 0.259 |
| CD4+ cell count/ul | 488.8±394.1 | 828.6±469.9 | 0.141 |
| Treg, % | 11.5±5.1 | 8.3±2.9 | 0.103 |
| Foxp3+Treg, % | 10.4±3.9 | 7.8±2.6/ | 0.127 |
| Teff, % | 89.6±3.6 | 91.6±2.8/ | 0.186 |

RPILD: rapidly progressive interstitial lung disease; CILD: chronic interstitial lung disease; OS: overall survival; MSAs: myositis-specific autoantibodies; MAAs: myositis-associated autoantibodies; ANA: anti-nuclear antibody; TIF-1γ : translation initiation factor-1γ ; MDA5: melanoma differentiation-associated 5; NXP2: nuclear matrix protein 2; SAE: small ubiquitin-like modifier enzyme; PM/Scl: polymyositis/scleroderma; RF: rheumatoid factor; AST: aspartate aminotransferase; LDH: lactate dehydrogenase; CK: creatine kinase; ESR: erythrocyte sedimentation rate; CRP: C-reactive protein; NK: natural killer cell; Treg: Regulatory T cells; Teff: effector T cells.

